# Robotic walking for recovery of functional capacity in individuals with incomplete spinal cord injury: A randomized pilot trial

**DOI:** 10.1101/2021.10.24.21265389

**Authors:** Claire Shackleton, Robert Evans, Sacha West, Wayne Derman, Yumna Albertus

**Affiliations:** Physical Activity, Lifestyle and Sport Research Centre (HPALS), Department of Human Biology, University of Cape Town, Cape Town, South Africa; Department of Sport Management, Cape Peninsula University of Technology, Cape Town, Western Cape, South Africa; Institute of Sport and Exercise Medicine, Faculty of Medicine and Health Sciences, Stellenbosch University, Tygerberg Campus, Cape Town, Western Cape, South Africa; IOC Research Center, South Africa

**Keywords:** Spinal cord injuries, rehabilitation, robotics, exoskeleton device, exercise therapy, gait, muscle strength

## Abstract

**Objective:** Activity-based Training (ABT) represents the current standard of neurological rehabilitation. Robotic Locomotor Training (RLT) is an innovative technique that aims to enhance rehabilitation outcomes, however, its efficacy in SCI rehabilitation, particularly within a low-middle income setting, is currently unclear. The primary aim of this study was to determine the feasibility of a locomotor training program within XX, in terms of recruitment, attendance, drop-out rates and safety. Secondary aims were to determine the effects of RLT compared to ABT on functional capacity in people with chronic SCI.

**Design:** Participants with chronic traumatic motor incomplete tetraplegia (n = 16) were recruited. Each intervention involved 60-minute sessions, 3x per week, over 24-weeks. RLT involved walking in the Ekso GT™ suit. ABT involved a combination of resistance, cardiovascular and weight-bearing exercise. Primary feasibility outcomes included recruitment rate, adherence rate, and adverse events. Validated tests were performed at baseline, 6, 12 and 24-weeks to assess the secondary outcomes of functional capacity.

**Results:** Out of 110 individuals who expressed interest in participating in the study, 17 initiated the program (recruitment rate = 15.4 %). Of these, 16 individuals completed the program (drop-out rate = 5.8 %) and attended sessions (attendance rate = 93.9%). There were no significant differences between the intervention groups for lower or upper extremity motor scores (UEMS effect size (ES) = 0.09; LEMS ES = 0.05), back strength (ES = 0.14) and abdominal strength (ES = 0.13) after training. However, both groups showed a significant increase of 2.00 points in UEMS and a significant increase in abdominal strength from pre- to post intervention. Only the RLT group showed a significant change in LEMS, with a mean increase of 3.00 [0.00; 16.5] points over time. Distance walked in the Functional Ambulatory Inventory (SCI-FAI) increased significantly (p = 0.02) over time only for the RLT group.

**Conclusions:** Recruitment, attrition and adherence rates of the intervention and outcomes justify a subsequent powered RCT comparing RLT to ABT as an effective rehabilitation tool for potentially improving functional strength and walking capacity in people with incomplete SCI.

## Introduction

After spinal cord injury (SCI), physiological changes secondary to partial paralysis often result in considerable neuromuscular fatigue, decreased muscle strength, physical and cardiovascular deconditioning and the reduced ability to voluntarily activate affected skeletal muscles.^1–3^ Muscular strength and conditioning play a vital role in improving functional performance, daily independence and psychological well-being for individuals with SCI.^2^ Sedentary behaviour and inactivity have also been linked to increased disability, and increased risk for cardiovascular disease and metabolic disorders.^2,4^ Therefore, there is a need to investigate rehabilitation interventions that promote recovery of functional capacity in people with chronic SCI.

The current global standard for neurological rehabilitation is that of Activity-based Training (ABT)^5–9^, with evidence that it improves multiple aspects of physical health and fitness after SCI.^10–12^ Research investigating the impact of robotic walking on physical parameters is growing, but due to a limited number of randomized control trials (RCTs), the effects on walking and functional capacity in people with SCI are unclear.^13^ Studies that focus on the effects of Robotic Locomotor Training (RLT) predominantly focus on treadmill and body-weight-support treadmill training, and not on over-ground RLT.^14–17^ However, it has been hypothesized that improvements from performing over-ground RLT would likely be similar to those found in other robotic interventions.^18–25^ Stronger evidence is needed to inform the development of evidence-based recommendations for RLT programs, particularly within low-middle income countries where out-patient rehabilitation strategies are extremely limited. Access to specialized healthcare services is not only a historic problem in XX, but it is further exacerbated by the lack of resources^26–28^, overstretched health systems and unavailability of affordable and accessible transport.^26,29^ Local medical facilities experience a high burden of trauma, and resources are primarily focused on optimising acute care rather than out-patient support structures.^30,31^ Furthermore, robotic suits are not covered by medical aids and health insurance, therefore, these advanced technologies are beyond many people’s reach. The high cost of robot devices raises the question of its efficiency in comparison with other training strategies.^32^

Thus, this study aimed at filling this knowledge gap by performing a pilot study in support of a larger-scaled RCT. The primary aim of the present study was to investigate the feasibility and safety of a RTL program with a robotic exoskeleton, compared to ABT, offered to individuals with chronic SCI. Specifically, the intent of the present study was to determine the recruitment, attendance, drop-out rates, and the adverse events of delivering an exoskeleton rehabilitation program in XX. Furthermore, we aimed to investigate the effect of RLT in comparison to ABT on functional capacity outcomes in a low-middle income country.

## Materials and Methods

### Study Design

The comprehensive protocol of this pilot randomized controlled trial has been registered on the XX Clinical Trials Registry (XXCTR201608001647143). Each participant provided written informed consent prior to the study. Randomization and group allocation (assigned via computer generation) was performed by the project manager after participants completed pre-intervention screening. This study was approved by the Faculty of Health Sciences Human Research Ethics Committee (HREC 718/2017) and was performed in accordance with the principles of the Declaration of Helsinki, ICH Good Clinical Practice (GCP), and the laws of XX. A post-trial care period of three months was implemented after participants completed the intervention, with continued access to rehabilitation equipment and medical professionals provided.

### Participants

The recruitment process is provided in detail in Figure 1 and spanned a period of 18 months. A total of 17 participants from the XX region in XX, with chronic (>1 year) SCI were recruited and randomly assigned to the RLT or ABT intervention groups. Inclusion criteria were chronic (>1 year), traumatic tetraplegia, individuals 18-65 years, motor incomplete injury (AIS C, D), with a neurological level of injury (NLI) between C1-C8 (tetraplegia), reliant upon a wheelchair as the primary mode of mobility, sufficient anthropometrics and range of motion (ROM) to achieve a normal, reciprocal gait pattern within the Ekso GT™ suit, were medically stable and cleared by a physician for full weight-bearing locomotor training, including 15-minute standing frame trial to assess standing tolerance.

**Figure 1:**
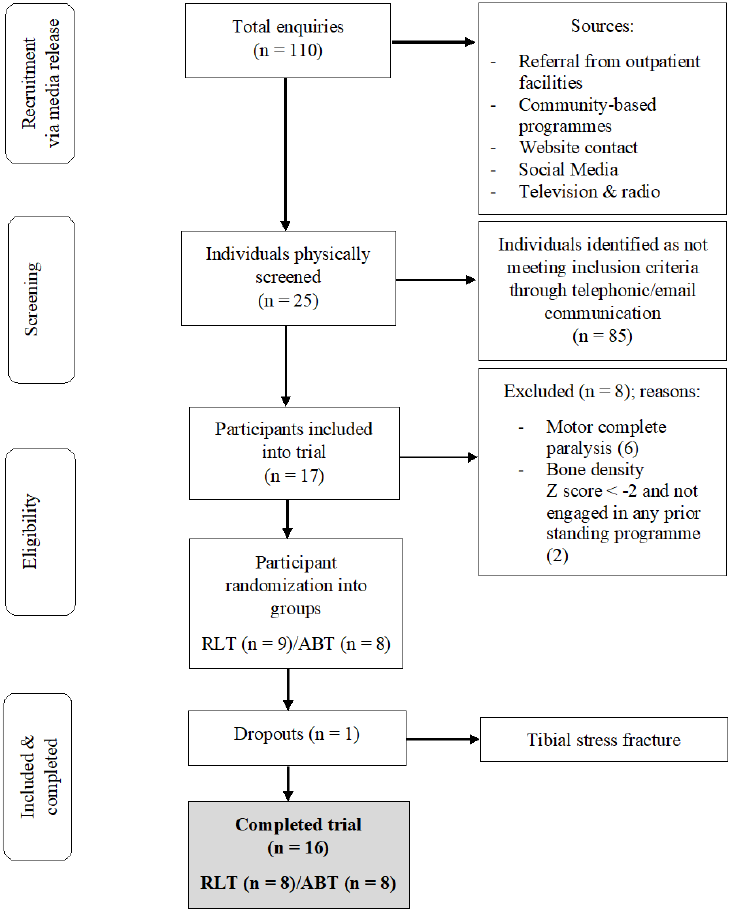
CONSORT flow chart of the recruitment process of participants into the trial.

Exclusion criteria included: non-traumatic SCI, had trained in a robotic exoskeleton in the past 12-months or were currently performing any other form of locomotor training, Modified Ashworth Scale (MAS) = 4 in any of the lower extremity joints, skin integrity issues in areas that contact the device, pregnancy, severe osteoporosis, any medical issue that in the opinion of the investigating team precluded full weight-bearing locomotor training, including but not limited to: heart or respiratory comorbidity, spinal instability, acute deep vein thrombosis (DVT) with activity restrictions, severe, recurrent autonomic dysreflexia (AD) requiring medical intervention, heterotopic ossification (HO) in the lower extremities resulting in ROM restrictions at the hips or knees, any medical issue that in the opinion of the investigating team would affect participant safety either due to cognitive deficits/impulsivity, intolerance to mild exercise or other factors, any issue that in the opinion of the investigating team would confound results such as a concurrent neurological injury or disorder (other than SCI).

### Interventions

The exercise intervention consisted of 24 weeks of supervised RLT and ABT. Both interventions consisted of three sessions per week, 60-minutes each, for 24-weeks and were overseen by trained healthcare professionals. RLT involved solely walking in an Ekso^®^ GT Variable Assist Model exoskeleton (Ekso Bionics, Richmond, CA, US), which has been used in previous research.^33,34^ Intensity levels were determined by the attending therapist and ranged from standing and walking time of 10 to 50 minutes and between 50 and 1800 steps taken. ABT consisted of a combination of resistance, cardiovascular, and flexibility training in various positions. Gait retraining, without a treadmill or robotic assistance, was also performed in the ABT group. Upper and lower body resistance training was performed using bodyweight exercises and various apparatus, including bands, wrist weights, dumbbells, and cables. The approximate standardized time allocation for each ABT session was as follows: warm-up and mobility (5 min), resistance training (20 - 30 min), and cardiovascular training (20 – 30 min). Five minutes were allocated for transfers and the setting up of various apparatus. Participants were monitored using the PARA-SCI tool and advised not to change their physical activity habits or dietary habits outside of the trial.

### Testing procedures

#### Feasibility outcomes

Recruitment was examined by recording the number of participants approached, screened for eligibility, and randomly assigned, as well as the reasons for exclusions. Adherence was determined by the number of participants who participated in the intervention, their attendance to scheduled sessions with reasons for dropouts and missed sessions. Adverse events to the intervention were monitored and recorded.

#### Functional capacity outcomes

The design of this pilot clinical evaluation comprised pre, 6 weeks, 12 weeks, and post (24 weeks) assessments of the intervention effect on functional outcomes. Specific methods pertaining to the individual functional outcomes are provided below.

### Handgrip strength

Handgrip strength of both hands was measured using a hand-held dynamometer (Camry Electric Hand Dynamometer Model EH101) to measure the maximum isometric strength of the hand and forearm muscles.^35^ The participants were encouraged with verbal cueing to provide a maximum effort.

### International Standards for Neurological Classification of Spinal Cord Injury (ISNCSCI) impairment scale

The International Standards for Neurological Classification of Spinal Cord Injury (ISNCSCI) involved an upper body and lower body motor examination to determine the neurological level of injury and whether the spinal injury was complete or incomplete.^36,37^

### Isometric dynamometry

The peak isometric force of abdominal flexion (rectus abdominus) and back extension (erector spinae and latissimus dorsi) were measured using the Biodex dynamometer (System3). Dynamometry set-up was standardized for each participant according to abdominal and leg length, with the dynamometric axis positioned at the third lumbar vertebral body to minimize error with repeated measures.^38^ The isometric strength tests included three maximal voluntary contractions (MVCs) of five seconds each, separated by two-minute rest intervals. During the MVCs, participants were verbally motivated and received post contraction feedback on the level of force generated to encourage them to reach their full potential in a subsequent attempt. MVCs were performed for abdominal flexion and back extension, with 10-minute rest period between the two protocols.

### 6-Minute Arm Ergometry Test (6MAT) and Rating of Perceived Exertion (RPE)

The 6-minute Arm Ergometry Test (6MAT)^39^ involved six minutes of submaximal exercise on a standard arm cycle ergometer at a constant power output. The aim was to attain a steady heart rate of 60% - 70% of age-predicted maximum heart rate. Set up of the arm ergometer was standardised for each participant according to their ergometer height and hand position. The Borg Rating of Perceived Exertion (RPE) and the distance covered was recorded on completion of the test.

### Spinal Cord Injury Functional Ambulatory Inventory (SCI-FAI)

The SCI-FAI was an observational gait assessment in which the participants performed a two-minute walk test with one of various levels of assistance (parallel bars, walker, crutch, or cane) whilst being video recorded (1-minute lateral view, 1-minute anterior view).^40^ A blinded independent assessor retrospectively scored the video footage. Outcome measures of this test included the distance walked, the technique score and the device score.

## Statistical analyses

All data were analysed using statistical software (R, R Core Team, Auckland, New Zealand and Prism 8, GraphPad Software Inc, California, USA). Normality was assessed using the Shapiro-Wilk test. Linear mixed effect models assessed continuous responses which were measured at four time points (0, 6, 12 and 24-weeks). To account for the within subject association between repeated measures, subject specific random effects were included (modelled coefficient p-values and 95% CIs). Response profiles were illustrated using plots of means and half-width 95% confidence intervals (CI) for observed data. Mann Whitney U and Wilcoxon Signed Rank tests were used to determine the group and time main effects for the non-parametric outcomes (motor scores). Significance was accepted at a p ≤ 0.05. Magnitude-based inferences of change (effect sizes) were calculated according to Cohen’s d^41^ for parametric data and the Mann Whitney U for non-parametric independent samples (r = z/√N).^42^ Effect size estimates of zero denote no effect, whereas ranges from 0.2 - 0.5, 0.5 - 0.8 and > 0.8 represent small, medium and large effects, respectively.^41^ The sample size determination for this trial has previously been reported on an additional primary outcome measuring cardiovascular outcomes.^43^ Whitehead et al suggested that a pilot sample size of 10 per treatment arm be reached for large effect sizes greater than 0.8.^44^ Due to eight participants in each group, we aimed for larger effect sizes for clinical significance.

## Results

### Participant characteristics

A total of 16 participants, aged 19 – 60 years (mean ± SD: 38.4 ± 14.3), completed the trial (Table 1). The RLT and ABT groups were matched at baseline for age and time since injury. Motor vehicle accidents accounted for 63% of injury aetiology in both groups, whilst stabbing, gunshot, rugby, motorcycle, mountain bicycle and diving accounted for 12.5% each.

**Table 1:** Participant characteristics of the Robotic Locomotor Training and Activity-based Training groups.

| Group | Participant | Age range (years) | Time since injury range (years) | Neurological level of injury | AIS category | Aetiology | Sex |
| --- | --- | --- | --- | --- | --- | --- | --- |
| RLT | 1 | 26 - 30 | 6 - 10 | Low cervical | D | Stabbing | Male |
|  | 2 | 31 - 35 | 11 - 15 | Low cervical | C | MVA | Male |
|  | 3 | 31 - 35 | 1 - 5 | Low cervical | D | MVA | Male |
|  | 4 | 46 - 50 | 26 - 30 | High cervical | D | Gunshot | Male |
|  | 5 | 51 - 55 | 1 - 5 | Low cervical | D | MVA | Male |
|  | 6 | 41 - 45 | 21 - 25 | Low cervical | C | MVA | Male |
|  | 7 | 56 - 60 | 11 - 15 | High cervical | C | MVA | Male |
|  | 8 | 31 - 35 | 11 - 15 | Low cervical | C | Sport - Rugby | Male |
|  | Average | 40.5 ± 11.2 | 13.8 ± 8.2 |  |  |  |  |
| ABT | 9 | 26 - 30 | 1 - 5 | Low cervical | C | MVA | Male |
|  | 10 | 46 - 50 | 16 - 20 | Low cervical | D | MVA | Female |
|  | 11 | 46 - 50 | 6 - 10 | Low cervical | D | MVA | Male |
|  | 12 | 15 - 20 | 1 - 5 | Low cervical | C | MVA | Male |
|  | 13 | 46 - 50 | 1 - 5 | High cervical | D | Motorcycle | Male |
|  | 14 | 26 - 30 | 6 - 10 | Low cervical | C | MVA | Male |
|  | 15 | 56 - 60 | 1 - 5 | Low cervical | C | Mountain bike | Male |
|  | 16 | 26 - 30 | 11 - 15 | High cervical | C | Diving | Male |
|  | Average | 38.4 ± 14.3 | 7.3 ± 6.4 |  |  |  |  |

*RLT*: Robotic Locomotor Training (n = 8); *ABT*: Activity-based Training (n = 8); *MVA*: motor vehicle accident. Values quoted as mean ± SD. No statistically significant difference between groups for age (p = 0.74) and time since injury (p = 0.10).

### Feasibility outcomes

Figure 1 shows the CONSORT flow diagram of participant recruitment and retention. There were 110 enquiries with 25 individuals who were screened for eligibility. Eight individuals did not meet the study criteria, with reasons provided in Figure 1. The other 17 individuals were enrolled and began the intervention (n = 8 RLT, n = 8 ABT). Participants had an average adherence of 93.9 ± 6.2% (67 out of 72 sessions) of all available sessions with no statistical difference in the adherence rate between groups. The participant with the lowest adherence achieved 83.3%, whilst three participants achieved a 100% adherence rate. Reasons for missing sessions (Fig. 2) were due to pathophysiological reasons such as bowel complications, urinary tract infections, baclofen pump complications and skin abrasions were localised to a few participants but occurred repetitively throughout the trial. Logistical reasons such as unreliable transport, work, personal and study commitments were more widely spread across participants. Only one session was interrupted by an exoskeleton malfunction. Adverse reactions were monitored to inform about acceptability and appropriateness of interventions for individuals with SCI in future studies. One participant discontinued the intervention after being enrolled in the RLT group for three weeks. Persistent right leg weakness necessitated a magnetic resonance imaging study (MRI) which provided images consistent with the diagnosis of a tibial stress fracture. Only baseline measures had been recorded for the participant which may have been confounded by an existing stress fracture. Thus, the participant was excluded from all analyses and received treatment for the fracture outside of the trial protocol. No other adverse events or negative side effects were reported.

**Figure 2:**
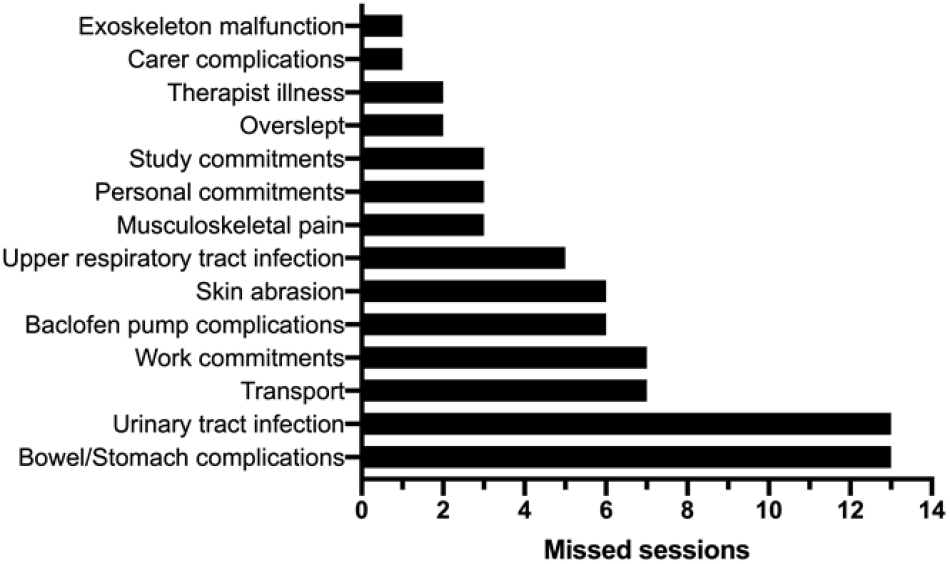
Reasons for missed rehabilitation sessions during the 24-week intervention. Notes: Overall, 72 sessions missed of 1152 total – 6.25%.

### Functional capacity outcomes

#### Strength capacity

There were no significant differences between groups for left (p = 0.75; ES = 0.16) and right handgrip strength (p = 0.30; ES = 0.61), respectively, nor for lower extremity motor scores (LEMS) (p = 0.86; ES = 0.05) (Table 2). However, only the RLT group showed a significant increase in LEMS from pre (16.00 ± 11.00) to post intervention (19.00 ± 11.00) (p < 0.05). There were no significant differences between the ABT and RLT groups for the upper extremity motor scores (UEMS) (p = 0.56; ES = 0.09). However, both groups showed a significant increase in UEMS from pre- to post (p = 0.03) intervention, with a mean difference of 2.00 points, respectively post intervention. There were no significant group differences for back (p = 0.77; ES = 0.14) or abdominal muscle strength (p = 0.80; ES = 0.13). However, both groups had a significant change in abdominal strength from pre-to post intervention (p = 0.02), with a mean increase of 7.04 [0.00; 22.35] Nm and 9.84 [0.00; 22.01] Nm for the RLT and ABT group, respectively (Table 2).

**Table 2:** A summary of functional capacity characteristics between the Robotic Locomotor Training and Activity-based Training groups.

| *Functional outcomes* | **RLT** |  |  |  | **ABT** |  |  |  |  |  |
| --- | --- | --- | --- | --- | --- | --- | --- | --- | --- | --- |
| | *Pre* | *Post* | $\Delta$ [95% *CI*] | *p* | *Pre* | *Post* | $\Delta$ [95% *CI*] | *p* | *Group p* | *Effect size* |
| Right handgrip (kg) | 11.42 ± 14.24 | 11.74 ± 14.60 | 0.32 [0.00;15.79] | 0.77 | 4.26 ± 4.42 | 4.99 ± 5.33 | 0.73 [0.00; 5.98] | 0.27 | 0.30 | 0.61 |
| Left handgrip (kg) | 8.49 ± 13.40 | 8.85 ± 14.39 | 0.36 [0.00; 15.27] | 0.75 | 5.79 ± 5.99 | 7.09 ± 5.74 | 1.30 [0.00; 7.59] | 0.24 | 0.75 | 0.16 |
| UEMS | 31.00 ± 11.00 | 33.00 ± 10.00 | 2.00 [0.00; 13.27] | 0.03* | 28.00 ± 10.00 | 30.00 ± 10.00 | 2.00 [0.00; 12.72] | 0.03* | 0.56 | 0.09 |
| LEMS | 16.00 ± 11.00 | 19.00 ± 14.00 | 3.00 [0.00; 16.5] | 0.05* | 16.00 ± 13.00 | 18.00 ± 14.00 | 2.00 [0.00; 16.49] | 0.11 | 0.86 | 0.05 |
| Abdominal strength (Nm) | 11.75 ± 8.89 | 18.79 ± 18.14 | 7.04 [0.00; 22.35] | 0.02* | 10.98 ± 10.03 | 20.82 ± 12.54 | 9.84 [0.00; 22.01] | 0.02* | 0.80 | 0.13 |
| Back strength (Nm) | 44.14 ± 32.87 | 68.21 ± 76.52 | 24.07 [0.00; 87.22] | 0.25 | 47.19 ± 29.93 | 78.55 ± 66.16 | 31.36 [0.00; 86.42] | 0.15 | 0.77 | 0.14 |
| 6MAT distance (km) | 1.05 ± 0.66 | 1.11 ± 0.66 | 0.06 [0.00; 0.76] | 0.33 | 1.04 ± 0.61 | 1.18 ± 0.60 | 0.14 [0.00; 0.78] | 0.09 | 0.83 | 0.11 |
| 6MAT RPE | 15.25 ± 2.87 | 16.88 ± 2.17 | 1.63 [0.00; 4.35] | 0.09 | 14.88 ± 2.10 | 13.88 ± 3.80 | -1.00 [0.00; 4.29] | 0.49 | 0.08 | **0.97** |
| SCI-FAI distance (m) | 2.57 ± 5.18 | 3.54 ± 5.84 | 0.97 [0.00; 6.88] | 0.02* | 1.51 ± 2.91 | 7.74 ± 9.83 | 6.23 [0.00; 14.00] | 0.10 | 0.47 | 0.53 |
| SCI-FAI technique score | 5.00 ± 5.73 | 8.00 ± 6.74 | 3.00 [0.00; 9.71] | 0.08 | 6.38 ± 7.25 | 8.25 ± 8.89 | 1.87 [0.00; 10.81] | 0.09 | 0.75 | 0.03 |
| SCI-FAI device score | 2.50 ± 2.98 | 3.00 ± 2.83 | 0.50 [0.00; 3.61] | 0.56 | 3.50 ± 3.82 | 4.00 ± 4.41 | 0.50 [0.00; 5.30] | 0.35 | 0.93 | 0.27 |

*RLT*: Robotic Locomotor Training group (n = 8); *ABT*: Activity-based Training group (n = 8); *kg*; kilograms; *UEMS*: upper extremity motor score; *LEMS*: lower extremity motor score; *Nm*;
newton meters; *6MAT*: 6-Minute Arm Ergo Test; *km*; kilometres; *RPE*; rating of perceived exertion; *SCI-FAI*; Spinal Cord Injury Functional Ambulatory Index (n = 7 due to outlier in ABT
group); *m*: meters; *Pre*: week 0 measurement; *Post*: week 24 measurement. Data presented as mean ± SD; $\Delta$ (95% *CI*): mean difference ± 95% confidence interval. Significance accepted at $p \leq$
0.05. Effect sizes (ES) between ABT and RLT groups at week-24: Bold represents a large effect size.

#### Endurance capacity

There were no significant differences between the ABT and RLT groups for 6MAT distance (p = 0.83; ES = 0.11) or rating of perceived exertion (RPE) (p = 0.08). However, a large effect size between groups at week 24 (ES = 0.97), highlights the reduced RPE during the 6MAT in the ABT group compared to the RLT group (Table 2).

#### Walking capacity

There were no significant between-group differences over time for distance walked during the SCI-FAI test (p = 0.47; ES = 0.53). However, only the RLT group had a significant improvement in distance walked over time (p = 0.02), with an increase of 0.97 [0.00; 6.88] m. A single participant in the ABT group (participant 5) presented as an outlier by walking substantially further than all the other participants across all time points (Fig. 3). This outlier achieved a maximum walking distance of 50 m compared to the others who scored between 0 and 25 m during the interventions. Six (n = 4 ABT; n = 2 RLT) of the 16 participants were non-ambulatory from baseline and continued to be so for the length of the intervention. Two participants in the RLT group who were non-ambulatory at baseline, both managed to achieve an improved distance of 2.44 m and 0.82 m by week 24 (Fig. 3). SCI-FAI device score and technique score remained unchanged for both interventions over time (Table 2). However, a single participant in the ABT group progressed from walking in the parallel bars to using a walker frame by week 24.

**Figure 3:**
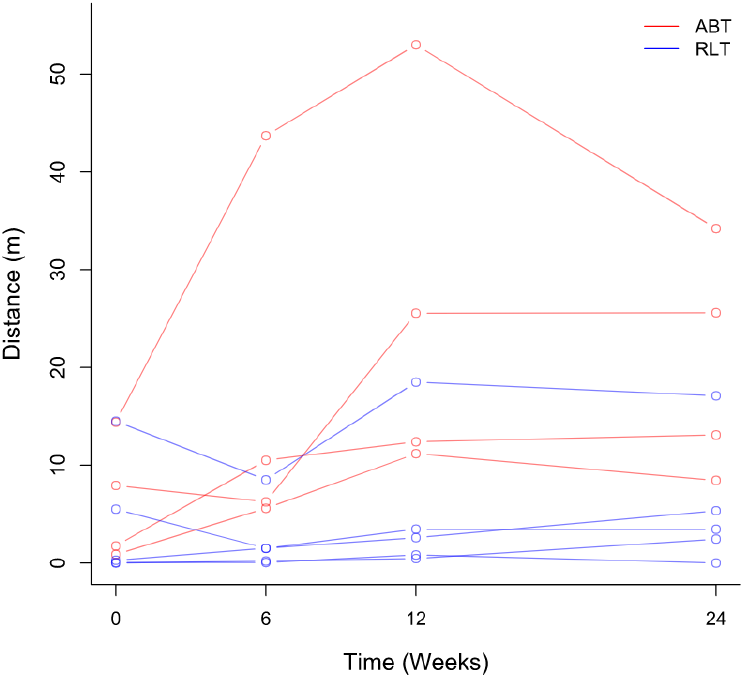
Individual participant (n = 16) SCI-FAI distance in the Robotic Locomotor Training and Activity-based Training groups over time. *RLT*: Robotic Locomotor Training (n = 8); *ABT*: Activity-based Training (n = 8); *Outlier:* single individual in the ABT group; *Distance:* measured in meters.

## Discussion

This pilot study demonstrates the feasibility and safety of delivering a robotic walking program for individuals with SCI to assess changes in functional capacity. There was a reasonable participant recruitment and strong adherence to training, with only one adverse event. This study provides preliminary insight on how functional recovery may be impacted by this type of training in a low-middle income context.

### Feasibility of protocol

The recruitment rate reached in this preliminary study (15.4%) was acceptable although it remains relatively low considering the number of potential participants (n = 110) who initially expressed their interest in participating in the proposed study. Moreover, considering that the study was conducted in a rehabilitation centre offering some of the only specialized SCI rehabilitation in the XX area of XX and that numerous strategies were implemented to overcome potential barriers (e.g., wide reaching recruitment implemented, telephone pre-screening interview to minimize the number of visits, free transport, free training sessions), a higher recruitment rate was anticipated (i.e., ≥ 50%). Nonetheless, this recruitment rate is similar to those reported in other feasibility studies investigating locomotor training programs with a robotic exoskeleton in individuals with complete or incomplete SCI.^45,46^ Recruiting individuals with SCI to clinical trials is challenging; however, exoskeletons are generally viewed positively for rehabilitation and health benefits, and the opportunity to use an exoskeleton may incentivize participation.^43,47^ The current study screened 25 individuals, but only 17 were eligible and enrolled in the study. Of the 17 participants who started the intervention, one withdrew from the study. Reasons for dropout were not due to the demands of the study, but due to an underlying, pre-existing adverse event, suggesting that the study protocol was well tolerated.

The attendance rate reached in the present preliminary study (93.9%) was excellent. The high attendance with respect to the scheduled training sessions confirms the commitment of the participants who engaged into the locomotor training program. This is further supported by the fact that only one participant dropped out of the program following an adverse event. Hence, a completion rate of 92.9% (n = 16 out of a total of 17 participants) was reached.

There was only one adverse event, in which a participant experienced a tibial stress fracture in week three of RLT. This fracture occurred even though the screening process was thoroughly completed by an experienced physician and the minimum Z-score BMD recommended by the manufacturer of the exoskeleton, was verified. In a comprehensive systematic review of RLT^48^, of the 18 studies that reported on adverse events, two reported small bone fractures, one of the talus^45^, and one of the calcaneus.^49^ These studies highlight that besides the potential positive effects of physical activity on bone structure, risks of developing adverse musculoskeletal effects require careful consideration.

### Functional capacity

There were no significant differences between the intervention groups for strength capacity outcomes. However, the participants in the RLT group experienced a significant increase in LEMS, with a mean change of 20% over time, compared to the non-significant change of 14% for the ABT group (Table 2). Buehner et al. (2012) support these findings where locomotor training induced a significant 21% improvement in LEMS (p < 0.01) after six months of RLT in adults with chronic incomplete SCI (ASIA C).^50^ Another study by Khan et al. (2019) involved 12 participants with chronic (> 1 year) SCI, who were trained in the ReWalk for 12 weeks. Two out of the three participants with motor incomplete injuries showed clinically meaningful improvements in LEMS.^51^ Increases in muscle strength are likely attributed to increased muscle fibre regeneration, increased muscle cross-sectional area and an increased ability to voluntarily activate affected skeletal muscles due to improved motor unit activation.^3,52,53^ However, motor recovery is dependent on the intensity, task specificity and goal-orientated approach of the training method, with overload and specificity being key principles for success.^53–57^

As RLT was more ambulatory-specific and more repetitive compared to the ABT intervention, lower limb strength may have been targeted more effectively in this group. There were no between-group differences for UEMS in this study, but both the RLT and ABT group showed significant increases in UEMS over time. These changes in UEMS after RLT may be attributed to the active participation from the user to move and use a gait aid while maintaining standing balance in the exoskeleton. Upper body strength gain within the ABT group was likely due to the key principle of training functional muscle groups within this modality.^58^

Although there was no significant group difference in abdominal strength, there were significant changes from pre-to post intervention for both the RLT and ABT group. The effects of RLT on trunk strength are currently unknown, as to our knowledge, no RLT studies have reported on these outcomes. However, RLT requires active participation from the user to shift their centre of mass forward and laterally in order to trigger the initiation of each step and to maintain an erect posture.^59,60^ Thus, the alternating weight-shifting and standing balance required in the exoskeleton, could help build and re-train the trunk musculature in people with SCI.^60^ Furthermore, dynamic trunk control is essential for stability during sitting and to perform other daily functional activities.^61^ Therefore, participants in the ABT group may have also adequately engaged the core muscles during the intervention to maintain postural stability and standing balance.

This study showed that after 24-weeks of ABT training, there was a trend (p = 0.09) towards a 13.5% improvement in 6MAT distance. Improved arm ergometry distance relies on both an increase in muscle strength as well as muscle endurance in order to produce force over multiple repetitions for the required six minutes.^2^ Comparable improvements in endurance capacity were reported by Jacobs et al. (2001) who showed that individuals with paraplegia were able to improve their upper limb endurance by 29.7% after completing a circuit training exercise program for 12-weeks.^62^ A similar study found that individuals who participated in a program which consisted of ABT twice per week for nine weeks, showed a significant 81% increase in submaximal arm ergometer power compared to a control group that did not participate in exercise.^63^

There were no significant between-group differences for the SCI-FAI, but there was a significant improvement in distance walked in the SCI-FAI test for the RLT group, with an increase of 0.98 m from pre-to post intervention. Additionally, two RLT participants who were non-ambulatory at baseline, both became ambulatory after training, with achievements of 2.44 m and 0.82 m by 24-weeks, whereas the four non-ambulatory individuals in the ABT group remained so throughout the intervention. Studies of locomotor training have demonstrated that recovery of walking and balance function can occur for individuals months and even years after incomplete SCI.^64,65^ Restoration of motor function is based on the repeated execution of motor tasks inducing plasticity and functional and structural reorganization of neuronal circuits in the injured spinal cord.^66–68^ The learning effect of walking must also be considered for improvements in walking capacity, as functional activities can also be improved by learning and using compensatory strategies.^56^

### Limitations

The functional status of the same ASIA Impairment Scale (AIS) category SCI is highly variable, thus, alternatives to subject enrolment based on AIS category should be considered in future studies. Other methods to achieve homogeneity of the SCI participants have been suggested, including stratification by LEMS or by initial ability to walk.^69^ Another limitation was the need to standardize the ABT program for purposes of the clinical trial. In practice, ABT tends to be highly individualized based on functional abilities, exercise limitations and preferences of the participant. This degree of individualization in a clinical trial would lead to virtually uninterpretable results, thus, standardization of therapy was needed according to the study guidelines. Lastly, owing to a small sample size, statistical comparisons between groups should be interpreted with caution. These limitations need to be considered when designing future RCTs with larger samples.

## Conclusion

This pilot study confirms that larger clinical trials investigating the effects of RLT compared to ABT are feasible and relatively safe in individuals with SCI within a XX context. The preliminary findings from this study have shown RLT to be a promising but costly rehabilitative tool to improve ambulatory function and functional strength capacity over time. These results should be considered preliminary, but it is anticipated that they will stimulate interest in conducting future larger-scale RCTs investigating the efficacy of locomotor training programs in long-term manual wheelchair users in a low-middle income setting. There is a need to develop the infrastructure and knowledge in XX to adequately provide for the long-term needs of all individuals with SCI; focusing on innovative rehabilitation to promote optimal functioning and improve psychological well-being. This innovative rehabilitation may be costly, however, once efficacy is established, it will serve as a catalyst for the development of low-cost alternatives.

## Data Availability

All data produced in the present study are available upon reasonable request to the authors

## Acknowledgements

XX for assistance with the statistical analysis. XX for monitoring the psychological well-being of the participants involved in this trial. XX for the health screening of the participants involved in the trial. XX for student financial support. A loaner exoskeleton was provided by Ekso Bionics for the duration of the study. Ekso Bionics were not involved in the conceptualisation, design, data analysis, interpretation, or dissemination of the study’s results.

## Authorship confirmation statement

All authors have reviewed and approved the manuscript prior to submission.

CS was responsible for concept design, conducting tests, collecting and analysing data and writing of the publication.

RE was responsible for concept design, conducting tests, collecting data and editing the publication.

SW was responsible for concept design, co-supervising the study, supplying grant funding, conducting tests and editing the publication.

WD was responsible for concept design, co-supervising the study, statistical support and editing the publication

YA was responsible as principal investigator, primary supervisor of the study, concept design, supplying grant funding, conducting tests, collecting data, statistical support and editing the publication.

